# Application of Deep Learning Models into the Prediction of Interleukin-6 and -8 Cytokines in Sickle Cell Anemia Patients

**DOI:** 10.1101/2023.06.27.23291971

**Authors:** Dylan Nguyen, Lukas Abraham, Ashesh Amatya

## Abstract

Interleukin-6 (IL6) and Interleukin-8 (IL8) are cytokines related to general immune function, but within Sickle Cell Anemia (SCA) patients, their overproduction tends to cause auto-immune reactions. These vital cytokines engage in the pathophysiology of SCA, but the extent to which they are associated with the genetics of the disease requires further exploration. This research paper seeks to further the study of IL6 and IL8 in SCA patients as well as the possibilities to predict their presence in patients based on Haptoglobin alleles and various other hematological factors using artificial neural networks. This was done through a cross-sectional study of 60 sickle anemia patients and 74 healthy individuals who provided the basis for the data of this study. The deep learning model found a non-linear correlation between the Haptoglobin alleles and the production of IL6 and IL8, predicting their over presence in SCA patients with an accuracy of 90.9% and r-squared value of 0.88 based on the given inputs. The machine learning models built in this paper have the potential to accelerate the development of targeted treatments and diagnoses to those suffering from Sickle Cell Anemia and its specific immune complications.

## 1. Introduction

Sickle cell anemia (SCA) is a hereditary blood disorder that affects millions of people worldwide. It affects historically marginalized groups such as African Ameri-cans, where 1 in 500 African Americans carry the autosomal recessive mutation in addition to the 300,000 infants who are born every year with sickle cell anemia (Aziza & Kondamudi, 2022). It is caused by a mutation in the beta-globin gene, which leads to the production of abnormal hemoglobin molecules. The abnormal hemoglobin molecules can cause red blood cells to become stiff, sticky, and misshapen, leading to a range of symptoms, including pain, fatigue, and organ damage (Li, Xuejin, et al, 2017).

In the context of Sickle Hemoglobin (Hb S), it is caused by the aforementioned mutation of a single nucleotide (HBB glu(E)6Val(A); GAG>GTG; rs334; HBB:c.20A>T; MIM: #603903) in the beta globin gene (#603903 - sickle cell disease - OMIM, n.d.). The result leads to Hb S being polymerized as well as glutamic acid being interchanged by valine at position 6 of the beta globin chain. This polymerization leads to acute and chronic vascular occlusion that ultimately leads to acute chest syndrome, strikes, chronic hemolytic anemia, inflammation, cell adhesion, tissue hypoxia, organ ischemia, and tissue infarctions (Kato, Gladwin, & Steinberg, 2006).

In addition, it’s been reported that the level of cytokines in patients with SCA is higher than those of healthy patients. These include higher levels of Interleukin-1β, IL-6, IL-8, and tumor necrosis factor alpha (TNF-α) (Saylor et al., 1995; Lanaro et al., 2009) accession numbers. If the accession numbers have not yet been obtained at the time of These pro-inflammatory cytokines pose health risks to the human body as they cause chronic endothelium activation and adhesion of sickled red cells. This, in turn, causes constant local tissue ischemia and necro-sis that is often attributed to SCA.

The presence of Hb S and cytokines ties back together because the secretion of interleukins such as IL-6 and IL-10 is stimulated by haptoglobin (Hp), an acute phase protein that increases in production in response to inflammation. While this is how Hp plays a role in IL-6 and Il-8, it also binds itself to plasma Hb in a process known as hemolysis. This forms a soluble complex known as the Hb-Hp complex, effectively filtering Hb out of the plasma. This is demonstrated in figure 1, where Hemoglobin and Haptoglobin combine to form the complex. The Hp 1 allele binding to free Hb in particular has been studied fairly extensively (Guetta et al., 2007). Hp acts as the first line of defense against the effects of free Hb previously discussed. While this is just one allele of Hp, Hp is polymorphic, with Hp 1-1, Hp 2-1, and Hp 2-2 being the three possible genotypes of Hp in humans.

**Figure 1:**
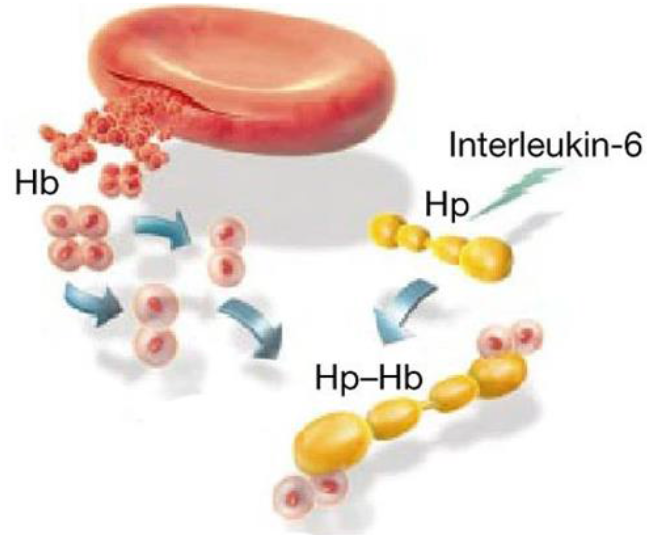
Model of Haptoglobin (Hp) binding with Hemoglobin (Hb) to form the Hp-Hb complex. Also demonstrates the secretion of Interleukin-6, but it does not depict the secretion of other previously mentioned cytokines. (Dennis, 2001)

The role of Hp genotype in regulating cytokine expression in patients with SCA, a disease marked by chronic and acute inflammation and hemolysis, has not been extensively researched, and present research has not found a conclusive correlation between the two. However, this project seeks to examine the link between different Hp polymorphisms and the pro-duction of cytokines IL-6 and IL-8 in SCA patients from Brazil compared to healthy individuals from the same area. This is done through building neural networks and other machine learning models that can accurately examine non-linear correlations between the two based on data sets of patients. Using artificial intelligence, this project hopes to build a model that can not only find a correlation through present data but find future correlations in other patients with SCA to expedite the process of diagnosis and development of treatments.

## 2. Material and Methods

### 2.1. Data Collection

The data collected in this study came from a pool of 134 individuals who participated in a separate study conducted by the Hereditary Anemia Outpatient Clinic of the Hematology and Blood Transfusion Unit, Universidade Federal de São Paulo (UNIFESP), Brazil. The subsequent data points were gathered by the authors of this paper in collaboration with UNIFESP, and the blood samples drawn were not drawn exclusively for this project. In addition, all blood drawings were done under a verified laboratory setting at UNIFESP. Of the 134 individuals whose data was used, 60 were diagnosed with SCA and the remaining 74 individuals were healthy. The 74 healthy individuals were age and gender matched Hb AA volunteers who came from approximately the same area as the 60 other individuals. Participants were all aged 13 and up, and informed consent was gathered from each individual, with parental consent being gathered from adolescent participants. Furthermore, the data collection has gotten the approval of UNIFESP’s own Research Ethics Committee.

To determine the plasma levels of IL-6 and IL-8 cytokines in the patient sample, blood samples were collected from the antecubital veins of the patients. This was done under a tourniquet with minimal pressure applied. The high sensitivity enzyme-linked immunosorbent assays (ELISA) were used to measure cytokine levels in plasma. The normal range for IL-6 was 2.2 to 12.1 pg/mL, with a limit of detection of 2.2 pg/mL. The normal range for IL-8 was 10.2 to 34.3 pg/mL, with a limit of detection of 0.8 pg/mL.

To extract DNA from buffy-coat samples, the stored samples were defrosted and subjected to red blood cell lysis with saponin. Leukocytes were washed with Solution A, followed by DNA extraction using Solution A without saponin and Solution B containing proteinase K. After incubation and purification, DNA was obtained using the Phenol:chloroform:isoamyl alcohol (25:24:1) method. The entire procedure was performed at the Molecular Biology Laboratory of UNIFESP, Brazil under professional and adult supervision. Haptoglobin (Hp) genotyping was carried out by a polymerase chain reaction (PCR) technique, which involved two protocols. The first was Protocol 1, which amplified the 1757 bp allele-1 specific sequence and the 3481 bp allele-2 specific sequence. The PCR products were then separated to confirm the presence of the haptoglobin genotype in the patients. This was done via the use of agarose gel electrophoresis.

### 2.2. Data Cleaning

The end values were analyzed using a variety of statistical tests. The average plasma levels of interleukins between cases and controls were analyzed using an independent sample t-test. The p-values calculated in Table 1 were done through three tests, that being the Chi-square test, Mann-Whitney test, and the independent sample t-test. The gender, Haptoglobin alleles, and Haptoglobin genotypes’ p-values were all calculated with the Chi-square test. The age in years’ p-value was calculated with the Mann-Whitney test. And finally, the Plasma IL levels’ p-value was calculated with the independent sample t-test.

**Table 1.** Demographics, haptoglobin genotypes, and plasma levels of IL-6 and IL-8 between Sickle Cell Anemia (SCA) patients and controls.

| Variables | SCA Group<br>(n=60) | Control Group<br>(n=74) | p-value |
| --- | --- | --- | --- |
| **Gender n (%)** |  |  |  |
| Male | 32 (53) | 39 (53) | 1.00 |
| Female | 29 (47) | 35 (47) |  |
| **Age in Years** |  |  |  |
| Median | 26.00 | 29.00 | 0.29 |
| Range | 15-50 | 17-60 |  |

| Plasma IL<br>(pg/ml)<br>mean ± SD | | | |
| --- | --- | --- | --- |
| IL-6 | 15.9 ± 9.1 | 4.4 ± 7.4 | <0.0001 |
| IL-8 | 20.5 ± 15.2 | 4.9 ± 7.4 | <0.0001 |
| Haptoglobin<br>Alleles<br>n (%) |  |  |  |
| Hp-1 | 54 (45.0) | 81 (54.7) | 0.113 |
| Hp-2 | 66 (55.0) | 67 (43.5) |  |
| Haptoglobin<br>Genotypes<br>n (%) |  |  |  |
| Hp 1-1 | 10 (16.7) | 25 (33.8) |  |
| Hp 2-1 | 34 (56.7) | 31 (41.9) | 0.07 |
| Hp 2-2 | 16 (26.6) | 18 (24.3) |  |

**Table 2.**
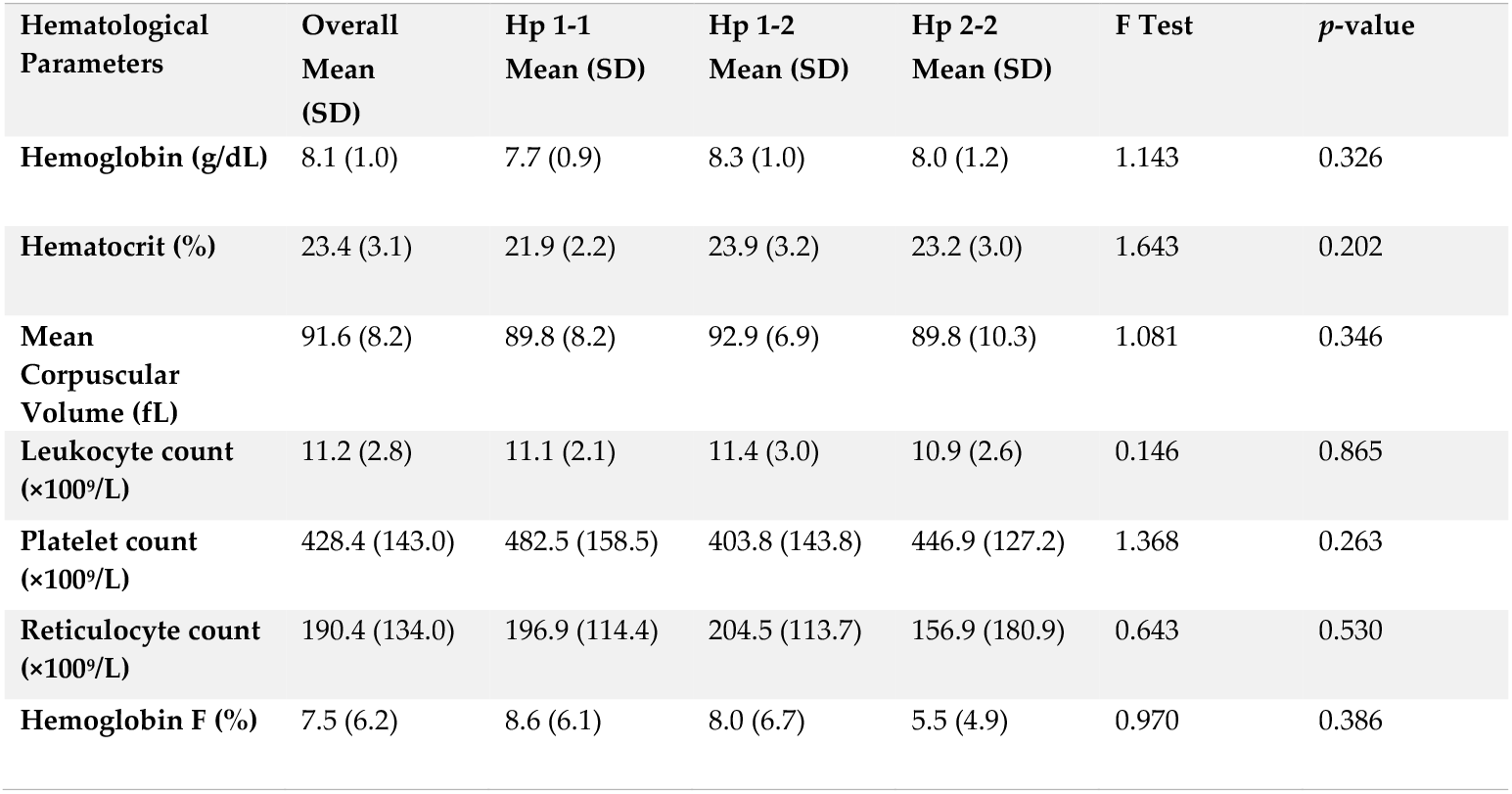
The hematological profile of the 60 patients who were diagnosed with Sickle Cell Disease

| Hematological Parameters | Overall Mean (SD) | Hp 1-1 Mean (SD) | Hp 1-2 Mean (SD) | Hp 2-2 Mean (SD) | F Test | *p*-value |
| --- | --- | --- | --- | --- | --- | --- |
| Hemoglobin (g/dL) | 8.1 (1.0) | 7.7 (0.9) | 8.3 (1.0) | 8.0 (1.2) | 1.143 | 0.326 |
| Hematocrit (%) | 23.4 (3.1) | 21.9 (2.2) | 23.9 (3.2) | 23.2 (3.0) | 1.643 | 0.202 |
| Mean Corpuscular Volume (fL) | 91.6 (8.2) | 89.8 (8.2) | 92.9 (6.9) | 89.8 (10.3) | 1.081 | 0.346 |
| Leukocyte count (×100<sup>9</sup>/L) | 11.2 (2.8) | 11.1 (2.1) | 11.4 (3.0) | 10.9 (2.6) | 0.146 | 0.865 |
| Platelet count (×100<sup>9</sup>/L) | 428.4 (143.0) | 482.5 (158.5) | 403.8 (143.8) | 446.9 (127.2) | 1.368 | 0.263 |
| Reticulocyte count (×100<sup>9</sup>/L) | 190.4 (134.0) | 196.9 (114.4) | 204.5 (113.7) | 156.9 (180.9) | 0.643 | 0.530 |
| Hemoglobin F (%) | 7.5 (6.2) | 8.6 (6.1) | 8.0 (6.7) | 5.5 (4.9) | 0.970 | 0.386 |

**Table 3.**
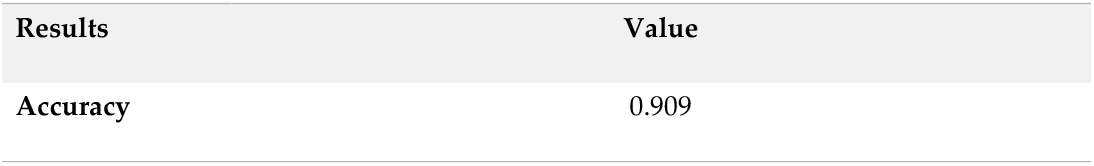
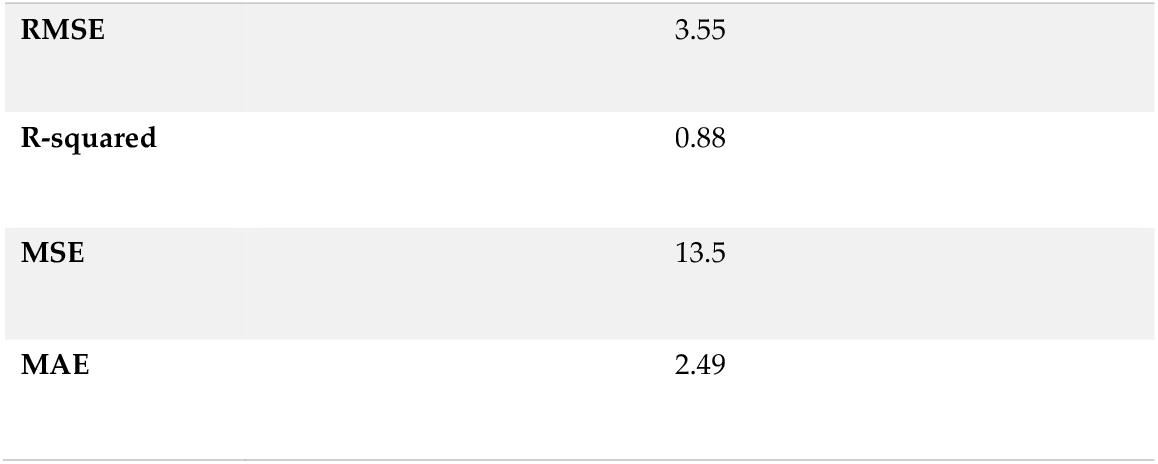
Various statistical averages from the ANN in predicting IL-6 and IL-8 Cytokines based on hematological factors and Haptoglobin alleles. These averages were over 100 trials of 500 epochs each.

## 3. Machine Learning Models

### 3.1. Data Collection

This project seeks to use machine learning to delve deeper and find real correlations between the Haptoglobin alleles and the levels of IL-6 and IL-8. This is done using a type of machine learning known as artificial neural networks. Artificial neural networks (ANNs) are a type of artificial intelligence that mimics human information processing into a program that can then be used for predictive purposes. These networks consist of algorithms that are modeled after the way human neurons work. ANNs use nodes that are similarly modeled after neurons in order to communicate signals with each other, and by linking multiple nodes together, they can recognize patterns and make predictions based on data. This application best suits the needs of predicting IL-6 and IL-8 levels based on subject data. This section seeks to describe the mathematics behind the ANNs used in this paper, including important concepts such as the activation function and learning rate.

### 3.2. Structure of an ANN

The specific ANN used in this project is a multi-layered perceptron (MLP) feedforward neural network. Its structure consists of an input layer, hidden layers, and output layers. In this case, the input layers are the various alleles and factors regarding the individuals, and the IL-6 and Il-8 cytokine levels will be the output layers. The hidden layer is where the variables of the input layers are passed through weights and bias in the nodes. The actual algorithm works through a process known as backpropagation, a diagram of which is shown in figure 2.

**Figure 2:**
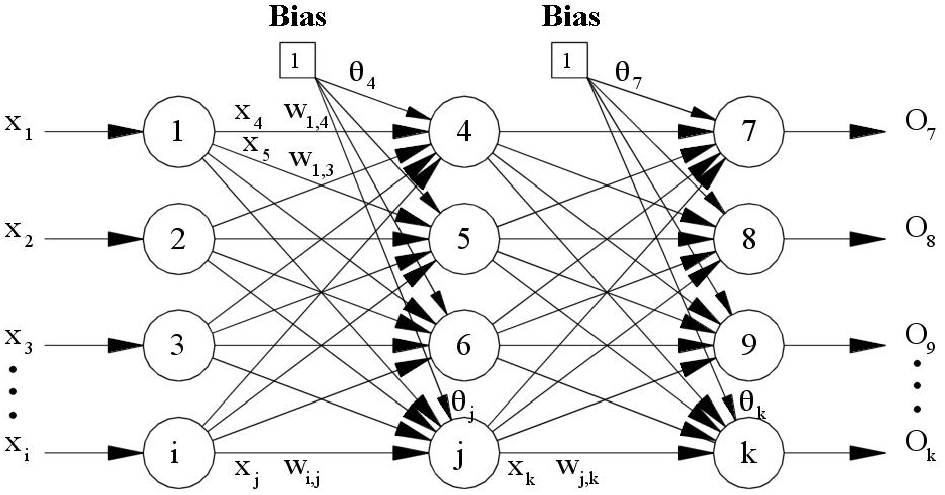
Backpropagation Neural Network with only one hidden layer. (Lu, 2000)

Backpropagation is defined as a neural network algorithm used for computing the gradient of the error with respect to the weights and updating the weights accordingly (Goodfellow, Bengio, & Courville, 2016). This is a supervised method of machine learning where the algorithm works by first making a forward pass through the network, computing the output of each neuron as a function of its inputs, weights, and bias. The difference between the predicted output and the actual output is then computed, and afterwards, the difference is mapped into a gradient that the algorithm will learn from to make a more accurate prediction for the second pass of input layers. This gradient is known as a gradient descent, and it’s shown in figure 3. (Mizutani, Dreyfus, & Nishio, 2000).

**Figure 3:**
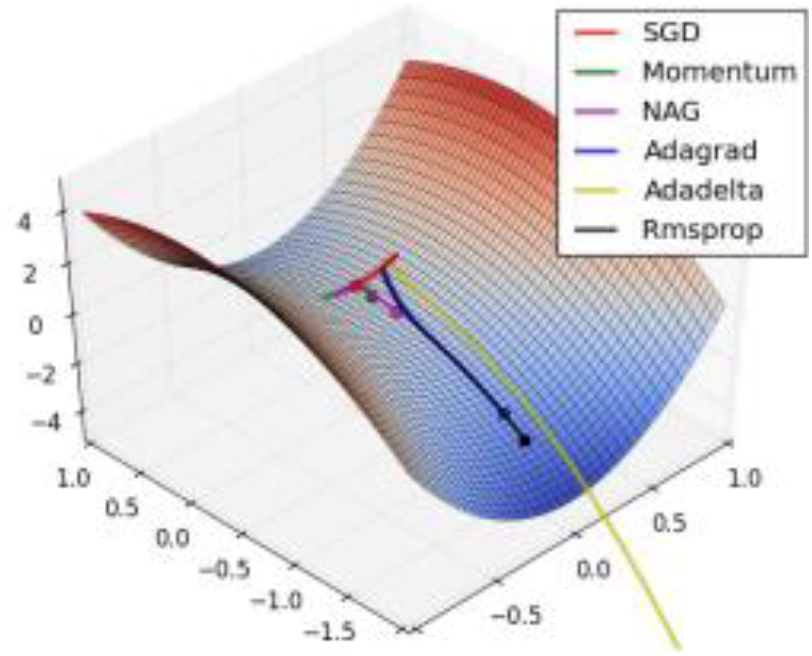
Three-dimensional representation of a Stochastic Gradient Descent model on saddle point. Courtesy of (Ruder, 2016)

**Figure 4:**
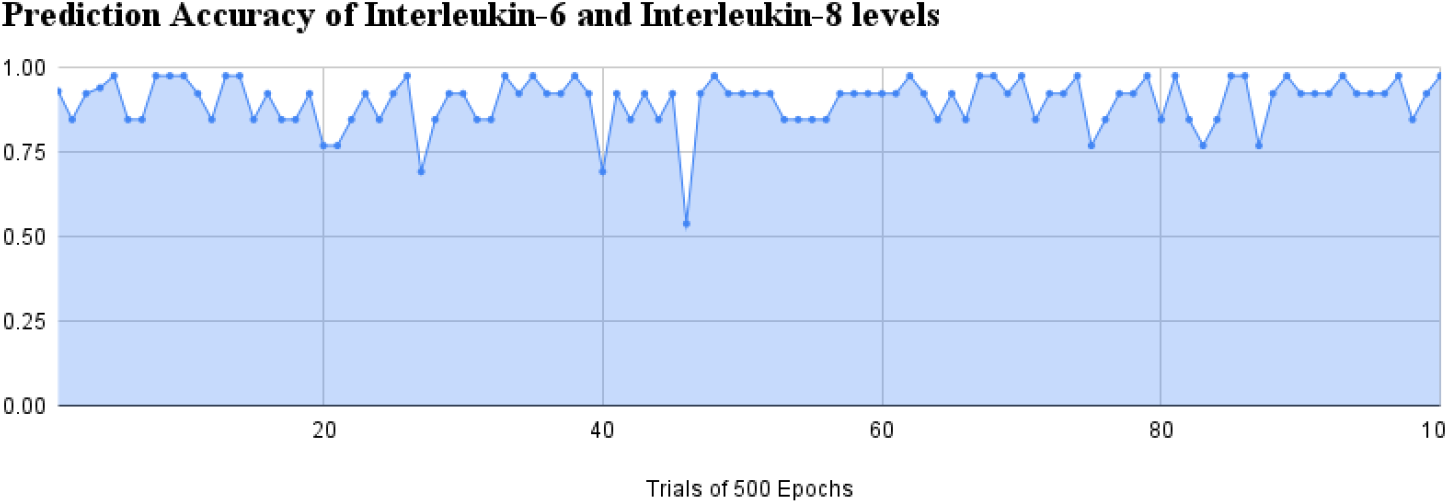
Chart displaying the accuracy of the artificial neural network over 100 trials of 500 epochs each.

When delving even deeper into how each ‘neuron’ works within the neural network, there’s a strong use of applied mathematics. For each neuron in the neural network, the weights in a neural network represent the strength of the connection between neurons. These will determine how much influence the input will have on the neuron’s output. In this case, the inputs will be the various alleles and values regarding a patient’s hematological profile. Drawing from a concept in linear algebra, the neural network takes the dot product of the input values while simultaneously adding weights and bias to shift the activation function left or right, allowing the end model to more accurate predict cytokine levels after each training epoch (Dasaradh, 2020).

### 3.3. Programming, Training, and Testing the Neural Network

The model within this research paper is built on Python using publicly available libraries such as TensorFlow, Sci-Kit Learn, pandas, and many others. The build environment for this neural network was Jupyter Notebooks, which is an open-source computing platform where all the Python code lives. It was integrated with Anaconda, which is a Python distribution / data discovery & analytics platform that allows this project to manage its packages better.

As for training the neural network, the total data was split into partitions of 70% and 30%. The 70% portion would be used to initially train the neural network and have it learn from the real-world inputs. The model was trained on 64 hidden nodes, and it was trained over 500 epochs. After much fine-tuning, 500 epochs allowed the model to get accustomed to the data of alleles and hematological profiles, but it wasn’t so much that the model started overfitting the data. The learning rate was set to 0.001 and the momentum was set to 0.9 as these values were found to give the neural network the best loss values.

Finally, there were a variety of statistical measures employed to measure the performance of the model in predicting IL-6 and IL-8 levels. Root mean squared error (RMSE), mean squared error (MSE), mean absolute error (MAE), coefficient of determination (Rsquared or R2), and finally, pure accuracy.

## 4. Results and Discussion

After testing data sets were applied, there are various trends that become evident. The test found a pure accuracy of 90.9%, a root mean squared error of 3.55, a mean squared error of 13.5, a mean absolute error of 2.49, and an R-squared value of 0.88. These results were derived from 100 trials of the algorithm that ran 500 epochs each. The squared, root, and absolute error all being on the lower side means the algorithm was able predict said cytokine levels with minimal error per trial. In addition, the pure accuracy and R-squared values tell much the same story.

The R-squared value being so close to 1 means the algorithm was able to effectively correlate the x and y variables. This indicates that 88.2% of the variability in the dependent variables (Interleukin-6 and Inter-leukin-8 cytokines) could be explained by the independent variable of the Haptoglobin alleles and hematological profiles.

When looking for trends across all 100 trials, there’s clear evidence that the little volatility between epochs of which the algorithm ran indicates the predictive power of the algorithm at work. While this could indicate underfitting or overfitting, this is also likely the work of a well-formed model that has understood the inputs well and is predicting the outputs and cytokine levels in a contained range.

## 5. Conclusions

Sickle Cell Diseases (SCD) such as Sickle Cell Anemia (SCA) affect over 20 million worldwide, with people of African, Mediterranean, and Middle Eastern descent being more prone to infection. As predicted, this project discovered a non-linear correlation between Haptoglobin alleles, hematological parameters, and the production of Interleukin-6 (IL-6) and Interleukin-8 (IL-8) in patients with SCA. The potential for machine learning as a whole to be applied to the field of Sickle Cell Anemia research is massive and cannot be understated. The research built today has the potential to accelerate some of the most promising technologies that currently exist to diagnose and treat SCD.

Methods of combating SCA, including blood transfusions and hydroxyurea, are beginning to be researched in depth, and this machine learning model can speed up the development of cures and treatments. Hydroxyurea, as well as other treatment methods, has been proven to increase mean MCV and hemoglobin levels (Steinberg, 2003; Platt, 2008). Matching this with the prediction of IL-6 and IL-8 levels, personalized doses and treatment periods of medication could be modeled uniquely for every patient depending on their specific hematological parameters to reach a healthy Interleukin level.

Due to the predictive nature of these neural networks built through this project, the ability to predict the phenotype of SCA during the first months of a patient’s life or even in the prenatal period (since alleles can be analyzed even before birth) could allow a more precise prognosis and individualized treatment for young patients (Steinberg & Adewoye, 2006). The results produced in this study clearly outline the viability of machine learning models to predict cytokine levels, specifically IL-6 and IL-8. Ultimately, the study’s results act as a net good for society.

## Data Availability

All data produced in the present study are available upon reasonable request to the authors.

## Author Contributions

Conceptualization, D.N.; methodology, D.N and L.A.; software, L.A and D.N.; validation, L.A., D.N. and A.A.; formal analysis, D.N and L.A.; investigation, D.N and L.A.; resources, D.N and L.A.; data curation D.N and L.A.; writing—original draft preparation, D.N and L.A; writing—review and editing, A.A.; visualization, D.N and L.A.; supervision, A.A.; project administration, A.A.; funding acquisition, A.A. All authors have read and agreed to the published version of the manuscript.

## Funding

This research received no external funding

## Informed Consent Statement

Not applicable

## Informed Consent Statement

Not applicable

## Data Availability Statement

Not applicable

## Acknowledgments

Bruna Spinella Pierrot-Gallo, Universidade Federal de São Paulo (UNIFESP), São Paulo, SP, Brazil.

## Conflicts of Interest

The authors declare no conflict of interest.

## Disclaimer/Publisher’s Note

^1^ indicates the authors contributed equally.

## Notes

### Competing Interest Statement

The authors have declared no competing interest.

### Funding Statement

This study did not receive any funding

### Author Declarations

Ethics committee/IRB of Universidade Federal de Sao Paulo (UNIFESP) gave ethical approval for this work.

## References

1. Sedrak, Aziza, and Noah P. Kondamudi. “Sickle Cell Disease.” StatsPearl, U.S. National Library of Medicine, 29 Aug. 2022 Author 1, A.; Author 2, B. Title of the chapter.

2. Li, Xuejin, et al. “Biomechanics and Biorheology of Red Blood Cells in Sickle Cell Anemia.” Journal of Biomechanics, U.S. National Library of Medicine, 4 Jan. 2017. Author 1, A.B.; Author 2, C. Title of Unpublished Work.

3. Kato, G. J., Gladwin, M. T., & Steinberg, M. H. (2006, November 7). Deconstructing sickle cell disease: Re-appraisal of the role of hemolysis in the development of clinical subphenotypes. Blood Reviews.

4. Taylor, S. C., Shacks, S. J., Mitchell, R. A., & Banks, A. (1995, December 15). Serum interleukin-6 levels in the steady state of sickle cell disease. Journal of interferon & cytokine research: the official journal of the International Society for Interferon and Cytokine Research.

5. Lanaro, C., Franco-Penteado, C. F., Albuqueque, D. M., Saad, S. T. O., Conran, N., & Costa, F. F. (2009, February). Altered levels of cytokines and inflammatory mediators in plasma and leukocytes of sickle cell anemia patients and effects of hydroxyurea therapy. Journal of Leukocyte Biology.

6. Guetta J, Strauss M, Levy NS, Fahoum L, Levy AP. (2006, July 3). Haptoglobin genotype modulates the balance of th1/th2 cytokines produced by macrophages exposed to free hemoglobin. Atherosclerosis.

7. Dennis, C. (2001). Hemoglobin Scavenger. Nature News. Retrieved February 20, 2023, from https://www.nature.com/arti-cles/35051680

8. Lu, W. (2000). Neural network model for distortion buckling behavior of cold-formed steel compression members. Helsinki University of Technology Laboratory of Steel Structures Publications, 16.

9. Goodfellow, I., Bengio, Y., & Courville, A. (2016). Deep learning. MIT Press.

10. Mizutani, E., Dreyfus, S., & Nishio, K. (2000). On derivation of MLP backpropagation from the Kelley-Bryson optimal-control gradient formula and its application. IEEE International Joint Conference on Neural Net-works.

11. Dasaradh. (2020, October 30). A Gentle Introduction to Math Behind Neural Networks.

12. Ruder, S. (2016, September 15). An overview of gradient descent optimization algorithms. ArXiv.

13. Steinberg, M. H., & Adewoye, A. H. (2006, May). Modifier genes and sickle cell anemia. Current Opinion in Hematology.

14. Steinberg, M. H. (2003, April 2). Effect of hydroxyurea on mortality and morbidity in adult sickle cell anemia. JAMA.

15. Platt, O. S. (2008, March 27). Hydroxyurea for the treatment of sickle cell anemia The New England Journal of Medicine.

